# The impact of the United States foreign aid freeze on HIV service delivery in PEPFAR-supported countries: a facility-level analysis of 2024–2025 programme data

**DOI:** 10.64898/2026.04.17.26351143

**Authors:** Brian Honermann, Anna Grimsrud, Elise Lankiewicz, Jennifer Sherwood, Greg Millett

## Abstract

**Introduction:** On January 20, 2025, the U.S. government froze foreign assistance including for PEPFAR, though a limited waiver for “life-saving” interventions was subsequently granted. PEPFAR’s 2025 monitoring results, released April 17, 2026, covered only quarter 4 while an earlier inadvertent release included all four quarters. Combining both data sets, we systematically assess facility-level programmatic performance and reporting trends to quantify service disruptions accounting for reporting discrepancies.

**Methods:** We categorized facilities by reporting continuity across Q1 2024 and Q4 2025 (e.g. continuous, intermittent, dropped, or new) and assessed changes in service delivery by the category of health facility for key HIV treatment, testing, PMTCT, and prevention programming. We additionally analyze changes in employed human resources for health (HRH) reported by PEPFAR.

**Results:** PEPFAR data included 31,746 facilities and community service sites. 71.3% were classified as continuous reporters, 16.9% intermittent reporters, 2.5% community services, 3.9% dropped in 2025, and 3.1% new in 2025. Total number of people accessing HIV treatment declined modestly by -0.3%, but differed by facility category. Continuous facilities saw a 0.5% increase in people on treatment, while intermittent facilities saw a -1.7% decrease. HIV testing declined -17%. HIV diagnoses declined -13% in continuous facilities, -35% in community services, and -29% in intermittent facilities. PMTCT infant testing and diagnoses declined by -6% and -12% in continuous facilities, respectively, and -60% and -31% in intermittent facilities, respectively. PrEP initiations declined -33%. Total direct service delivery HCWs reduced -62,541 (-24%)

**Conclusion:** These findings reveal substantial disruptions across PEPFAR service areas, with the steepest declines among intermittent and community-based delivery sites, alongside a 24% reduction in direct service delivery healthcare workers. As potentially the final data set PEPFAR will ever release, these findings represent a troubling inflection point. The dismantling of public data systems and accountability structures undermine progress and enable programmatic gaps to develop and go unnoticed that risk allowing HIV resurgence to occur over the coming years.

## Introduction

The United States (US) President’s Emergency Plan for AIDS Relief (PEPFAR), a bilateral program supporting HIV services in 55 countries, has been credited with saving more than 26 million lives since its inception in 2003.[1] While PEPFAR’s earliest successes centered on initiating PLHIV on ART, by 2024 the program had expanded well beyond treatment, including conducting more than 200,000 HIV tests daily, providing services to approximately 17,695 orphans and vulnerable children each day, and supporting a total of more than 700,000 people on pre-exposure prophylaxis (PrEP).[2,3] PEPFAR investments have been associated with both accelerated epidemic control and measurable health system strengthening in supported countries: new HIV infections fell 52% between 2010 and 2023 in PEPFAR-supported settings versus 39% globally, while domestic government health spending and health workforce capacity expanded at nearly double the rate seen in non-PEPFAR-supported countries over the same period.[1,4]

In early 2025, however, a series of changes to US foreign policy introduced significant alterations to the implementation of PEPFAR. On January 20, 2025 an executive order required a freeze on all foreign aid disbursements followed by a stop work order on January 24 stopped implementation of all foreign aid awards including PEPFAR, with minimal exceptions for humanitarian aid.[5,6] Following this period, a series of waivers allowed for the partial resumption of PEPFAR programming, including a PEPFAR-specific waiver that permitted a defined subset of HIV care, treatment, and prevention of mother-to-child transmission services to continue.[7] The subsequent months were characterized by considerable instability, including cycles of award cancellations and reinstatements, legal challenges to the freeze, and the permanent dissolution of USAID, which was one of two major PEPFAR implementing agencies. [8]

The full implications of the foreign aid review have been difficult to assess: the US government released no official list of terminated and active awards post-review so preliminary assessments relied on leaked lists that may have been incomplete or outdated. Assessments of these lists found 71% of HIV-focused USAID awards cancelled by number, while estimated cancellations accounted for only 24% of planned PEPFAR funding by value.[9–11] The downstream consequences of these funding disruptions have been characterized by a rapidly growing body of literature, though to date, primarily through modeling studies rather than observed programmatic data. Several studies modelled the potential impact of PEPFAR funding disruptions across a range of scenarios. Under a 90-day freeze, Hontelez et al. estimated 60,000–74,000 excess HIV deaths across seven sub-Saharan African countries;[12] another series of models estimated the projected impacts under full or near-full discontinuation with estimates across multiple modeling studies ranging from 4.4–10.75 million additional infections and up to 4.1 million additional deaths by 2030 above the status quo.[13,14]

Empirical studies have since supplemented modelling evidence, broadly demonstrating a period of initial severe disruption, followed by uneven recovery. Immediately post freeze, 71% of PEPFAR implementing partners reported cancelling at least one service category and 86% reported clients would lose ART access within a month.[15] As waivers were implemented, some clinical services, especially HIV treatment, resumed in many countries, but community-led services, PrEP, HIV testing, and key populations programming faced continued severe disruption. This unevenness is reflected in a number of assessments: the IeDEA consortium found 47% of clinics across 32 countries still reported care disruptions by mid-2025 while the UNAIDS country-level impact tracker found most severe impacts on community-level services but continued access to ART in many contexts.[16,17]

Critically, until April 10, 2026, PEPFAR Monitoring, Evaluation, and Reporting (MER) data for 2025, the most direct measure of PEPFAR programmatic performance, had not yet been made officially available for the full period from the major policy changes to date. The only available cross-country analysis of observed PEPFAR programmatic data relies on draft 2025 MER data obtained outside of official channels, and conducted only at the global level found substantial declines in HIV testing, viral load monitoring, and net ART coverage immediately post policy changes in early 2025, with partial but incomplete recovery by the end of September of the same year.[18] That assessment, however, was not a comprehensive assessment of the data available and only looked at aggregated results.

Using the newly available full MER dataset for this period, this analysis provides a systematic, facility-level assessment of PEPFAR programmatic performance and reporting trends during this period, offering a more complete picture of the impacts of the 2025 policy changes.

## Methods

### Data sources and study period

We used PEPFAR Monitoring, Evaluation, and Reporting (MER) data released on the PEPFAR development dashboard in January, 2026[18], alongside officially released data from April 17, 2026.

PEPFAR follows the U.S. government’s fiscal year (FY), with Q1 (October-December), Q2( January-March), Q3 (April-June), and Q4 (July-September). The major policy disruptions in early 2025 occurred during FY 2025 Q2. Officially released 2025 data only included Q4 data, whereas the earlier (unofficial) data set included all four quarters.

To assess the reliability of the unofficial dataset, we compared FY2025 Q4 results across both datasets at the sub-national and facility levels.

### Analytic approach and unit of analysis

Facility-level data were used to distinguish between true service delivery disruptions and apparent declines driven by loss of reporting when facilities ceased receiving PEPFAR support.

Because PEPFAR aggregates data only from actively supported facilities, higher-level aggregates cannot differentiate between service-level disruptions and reporting discontinuation. Notably, over 75% of health facilities reporting are government-owned, with remaining sites including faith-based, non-governmental organizational (NGO) -operated, and community outreach sites.

### Facility classification, data processing and definitions

Facilities were categorized into seven groups based on reporting patters across eight quarters (2024 Q1 - 2025 Q4):

- **Continuous facilities:** reported data in all 8 quarters;
- **Intermittent facilities:** reported in both Q4 2024 and Q4 2025 but had gaps in other quarters;
- **Community services:** records without facility codes (typically outreach programmes);
- **Dropped in 2025:** Facilities that reported data in Q4 2024, but reported no data as of Q4 2025;
- **New in 2025:** began reporting during 2025 with no prior data in 2024;
- **Previously dropped:** Facilities that ceased reporting data sometime before Q4 2024;
- **Other:** facilities not fitting the above categories.

Classification was based on the presence of any reported indicator, rather than indicator-specific reporting. TX_NET_NEW (Quarterly increase or decrease in patients on treatment) was excluded, as it is a derived variable rather than a directly reported measure.

Because PEPFAR data does not distinguish between the true zero values and missing data, the presence or absence of any reported value was used as a proxy for active reporting.

Community sites were assigned individual unique indicators at the lowest geographic level (generally a district) to provide a mechanism for counting them as distinct “sites”.

Facility identifiers were constructed by combining sub-national unit (SNU) identifiers to ensure uniqueness. Community service sites were assigned unique identifiers at the lowest geographic level (typically district).

### Outcome measures

We assessed changes between 2024 and 2025 across key HIV service delivery indicators:

- Number of people on treatment;
- HIV testing and diagnoses;
- Prevention of mother-to-child transmission (PMTCT) programming; and
- Pre-exposure prophylaxis (PrEP) initiations.

Analyses were conducted across all facilities categories, at the PEPFAR-wide level, and by country.

### Human resources for health analysis

We analysed PEPFAR Human Resources for Health (HRH) data to assess changes in workforce composition between 2024 and 2025. We excluded aggregate cadre data in 2024 as it was not directly comparable to 2025; and regional programme staff not assigned to specific countries.

These exclusions represented only 5.32% of total staff in 2024 and 0.08% of staff across the two years, respectively.

### Analysis

We report absolute and relative changes between 2024 and 2025. For intermittent facilities, quarter-to-quarter comparisons (Q4 2024 vs Q4 2025) were used where full-year data were incomplete.

## Results

### Changes in results from official and development site data

Q4 2025 data were only minimally different in the final official data set compared with the development site and were limited to entirely to South Africa affecting 132 facilities. Primarily these were facilities that had no reported results in the development site data. Full changes are available in the supplemental materials.

### Facility reporting patterns and classification

A total of 31,746 facilities and community service sites were included in the analysis (Figure 1). Most facilities (71.3%) reported continuously across all eight quarters from Q1 2024-Q4 2025, while 16.9% were classified as intermittent reporters. Smaller proportions of facilities were dropped in 2025 (3.9%) or newly reported in 2025 (3.1%), with community service sites comprising 2.5% of all reporting units.

**Figure 1:**
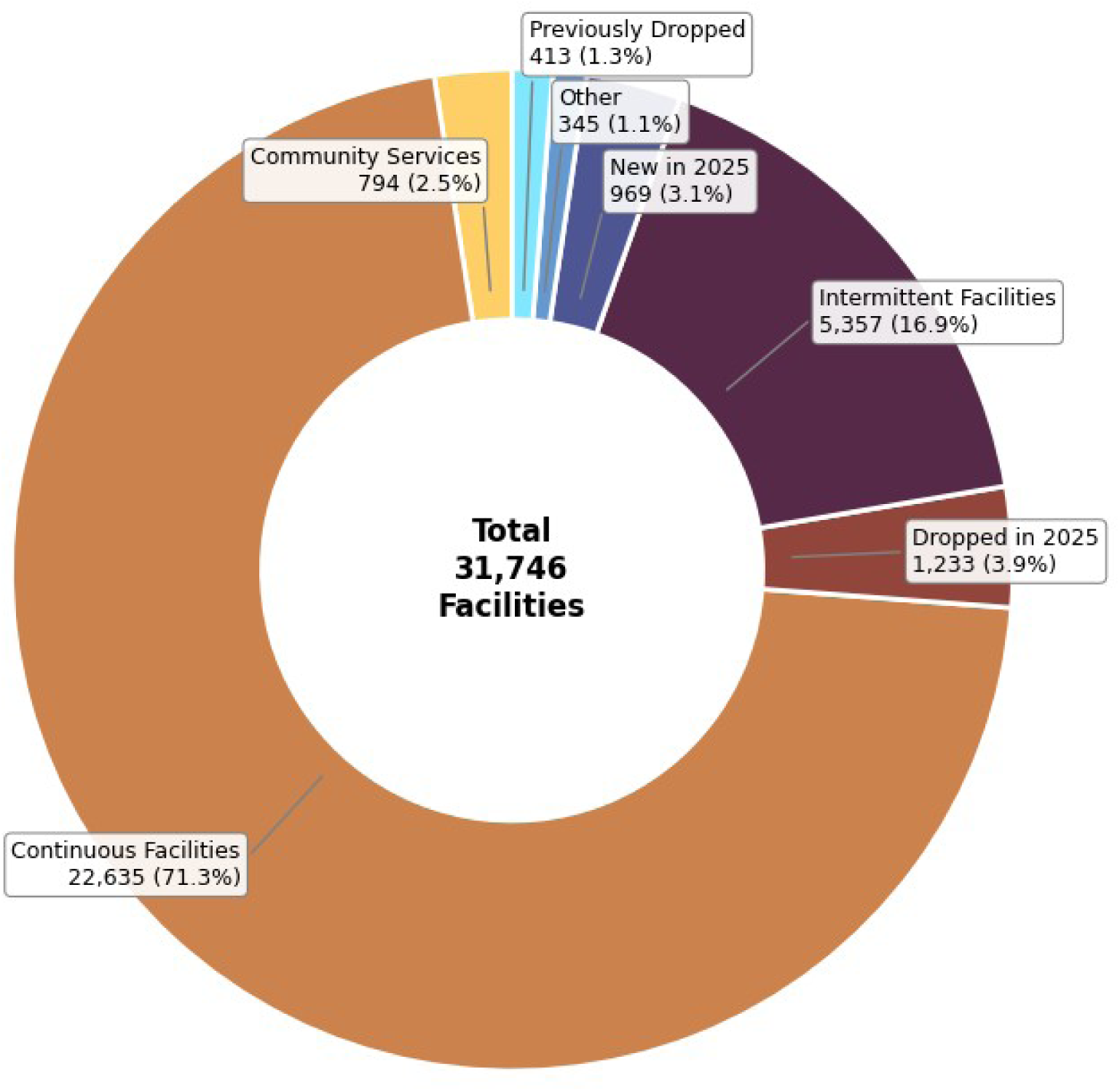
Continuous Facilities: Reported data every quarter from 2024Q1 - 2025Q4; Intermittent Facilities: Reported data in 2024Q4 and 2025Q4, but reported no data in one or more other quarters; Community Services: Sites in which the “Facility” code is blank; Dropped in 2025: Reported data in 2024Q4, but reported no data as of 2025Q4; New in 2025: Reported no data from 2024Q1-2024Q4, but began reporting sometime during 2025; Previously Dropped: Stopped reporting data sometime before 2024Q4; Other: Facilities that didn’t fall into any of the above categories.

Country-level variation in reporting patterns is show in Figure 2 (see Supplement 1 for full breakdowns).

**Figure 2:**
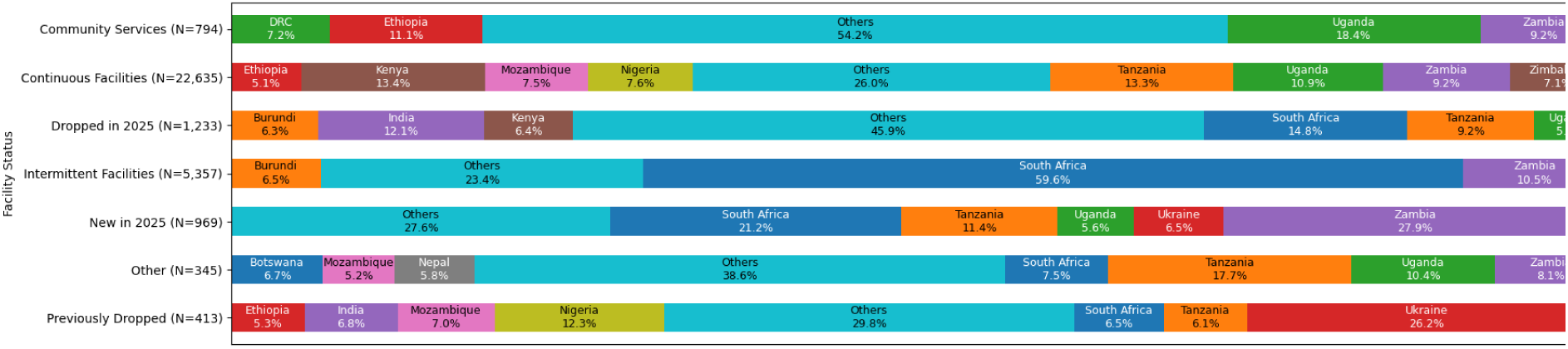
Facility Categories by Country Facility Count Contribution

South Africa was a noted outlier, with a large increase in facilities classified as Intermittent resulting in 59.6% of all such facilities. This is consistent with long-standing practice in South Africa that the National Department of Health contributes data to PEPFAR for all public clinics once annually rather than quarterly, while other implementing partners in South Africa continue to report quarterly. However, the number of facilities reporting on this basis in South Africa increased from 1,295 to 2,426 from 2024Q4 to 2025Q4, indicating a drop in direct implementing partner support in many parts of the country.

Burundi and Nicaragua had no facilities classified as continuous.

### Impact on the number of people accessing HIV treatment

Across all facilities, the total number of people accessing HIV treatment declined modestly by -65,064 (-0.3%) between Q4 2024 and Q4 2025 (Table 1). However, trends differed substantially by facility type. In Continuous facilities, the number of people accessing treatment grew by 0.5% (72,508), though growth was 48.6% lower than 2024 (Q1-Q4). Intermittent facilities, which are more likely to be representative of facilities that lost support of a PEPFAR IP, saw a decrease in the number of people accessing treatment (N=-74,607, -1.7%). Dropped facilities had supported 441,822 people in Q4 2024 with unknown status in 2025. New facilities supported 370,846 people on treatment by Q4 2025.

**Table 1:** HIV Treatment Indicators (2024Q1 - 2024Q4)

| HIV Treatment Initiations, Current, and Returned Patients by Facility Categorization (2024Q1-2025Q4) |  |  |  |  |  |  |  |  |  |  |  |  |
| --- | --- | --- | --- | --- | --- | --- | --- | --- | --- | --- | --- | --- |
|  | 2024 |  |  |  |  | 2025 |  |  |  |  | Increase/<br>Decrease | Change<br>(%) |
|  | Q1 | Q2 | Q3 | Q4 | Total | Q1 | Q2 | Q3 | Q4 | Total |  |  |
|  | Current on Treatment |  |  |  |  |  |  |  |  |  |  |  |
| Continuous Facilities | 15,527,759 | 15,475,614 | 15,503,356 | 15,659,944 | 15,659,944 | 15,664,481 | 15,318,402 | 15,478,305 | 15,732,452 | 15,732,452 | 72,508 | 0% |
| Dropped in 2025 | 355,159 | 363,280 | 366,059 | 441,822 | 441,822 | 96,771 | 34,195 | 12,593 |  | 0 | -441,822 | -100% |
| Intermittent Facilities | 3,175,515 | 3,256,067 | 3,294,542 | 4,298,080 | 4,298,080 | 3,290,765 | 174,841 | 1,312,991 | 4,223,473 | 4,223,473 | -74,607 | -2% |
| New in 2025 |  |  |  |  | 0 | 32,451 | 38,470 | 51,290 | 370,846 | 370,846 | 370,846 | inf% |
| Other | 7,635 | 2,049 | 3,424 |  | 0 | 7,320 | 9,503 | 7,943 | 8,011 | 8,011 | 8,011 | inf% |
| Previously Dropped | 58,907 | 55,015 | 48,040 |  | 0 |  |  |  |  | 0 | 0 |  |
| All Facilities | 19,124,975 | 19,152,025 | 19,215,421 | 20,399,846 | 20,399,846 | 19,091,788 | 15,575,411 | 16,863,122 | 20,334,782 | 20,334,782 | -65,064 | -0% |
|  | Newly Initiated on Treatment |  |  |  |  |  |  |  |  |  |  |  |
| Continuous Facilities | 318,943 | 334,559 | 311,293 | 308,375 | 1,273,170 | 284,209 | 248,496 | 266,635 | 273,877 | 1,073,217 | -199,953 | -16% |
| Dropped in 2025 | 10,032 | 9,854 | 9,714 | 16,656 | 46,256 | 3,513 | 714 | 233 |  | 4,460 | -41,796 | -90% |
| Intermittent Facilities | 58,179 | 67,215 | 59,790 | 138,497 | 323,681 | 35,020 | 5,998 | 21,286 | 99,175 | 161,479 | -162,202 | -50% |
| New in 2025 |  |  |  |  | 0 | 1,647 | 1,918 | 2,446 | 15,743 | 21,754 | 21,754 | inf% |
| Other | 333 | 98 | 83 |  | 514 | 265 | 241 | 267 | 314 | 1,087 | 573 | 111% |
| Previously Dropped | 1,488 | 1,302 | 699 |  | 3,489 |  |  |  |  | 0 | -3,489 | -100% |
| All Facilities | 388,975 | 413,028 | 381,579 | 463,528 | 1,647,110 | 324,654 | 257,367 | 290,867 | 389,109 | 1,261,997 | -385,113 | -23% |
|  | Patients Returned to Treatment |  |  |  |  |  |  |  |  |  |  |  |
| Continuous Facilities | 250,072 | 345,361 | 316,300 | 306,224 | 1,217,957 | 225,161 | 251,754 | 329,266 | 298,448 | 1,104,629 | -113,328 | -9% |
| Dropped in 2025 | 6,481 | 7,154 | 7,390 | 7,155 | 28,180 | 1,221 | 319 | 591 |  | 2,131 | -26,049 | -92% |
| Intermittent Facilities | 91,485 | 115,867 | 107,908 | 105,119 | 420,379 | 47,103 | 3,516 | 42,794 | 49,989 | 143,402 | -276,977 | -66% |
| New in 2025 |  |  |  |  | 0 | 840 | 1,363 | 1,521 | 2,073 | 5,797 | 5,797 | inf% |
| Other | 272 | 34 | 110 |  | 416 | 167 | 266 | 382 | 917 | 1,732 | 1,316 | 316% |
| Previously Dropped | 1,465 | 1,694 | 1,172 |  | 4,331 |  |  |  |  | 0 | -4,331 | -100% |
| All Facilities | 349,775 | 470,110 | 432,880 | 418,498 | 1,671,263 | 274,492 | 257,218 | 374,554 | 351,427 | 1,257,691 | -413,572 | -25% |

The impact on the number of people accessing treatment by country in continuous facilities is shown in Figure 3.

**Figure 3:**
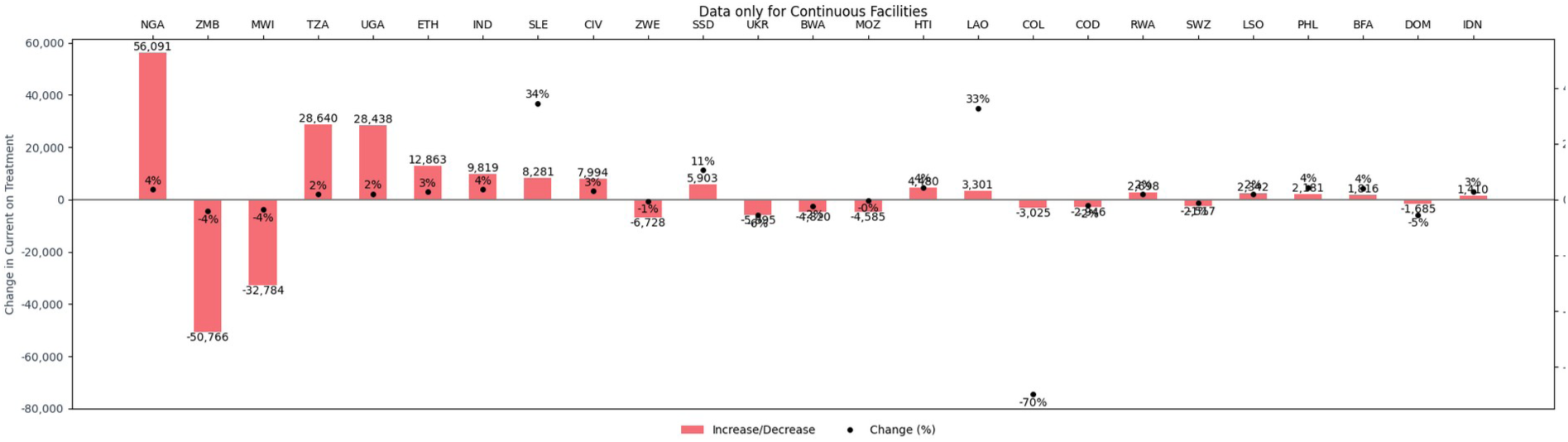
Change in People on Treatment by Country - 25 countries with greatest numerical change

Geographical analysis showed limited overlap between dropped and newly added facilities. In the 221 geographies where 701 treatment sites were dropped in 2025, only 105 geographies (48%) added new facilities. In areas where treatment sites were lost, the overall number of people accessing ART decreased -2% (N=-201,087). In the 105 geographies with both dropped and new sites, 110,318 people were added to treatment (2%). In the 116 geographies with dropped sites and no new facilities added, treatment rolls decreased by -311,405 (-6%). In geographies with new facilities but no dropped sites, 34,313 (2%) were added to treatment.

This finding is exemplified in Thailand where in 2024 53,329 people were supported on treatment in 29 sites. In 2025, 23 sites were dropped in 2025 and 14 sites were added, and the number of people supported on treatment increased 389% (N=207,652).

### Impact on the number of people tested, diagnosed, and initiation on treatment

PEPFAR-supported HIV testing declined by -14.0 million tests between 2024 and 2025 (Table 2).

**Table 2:**
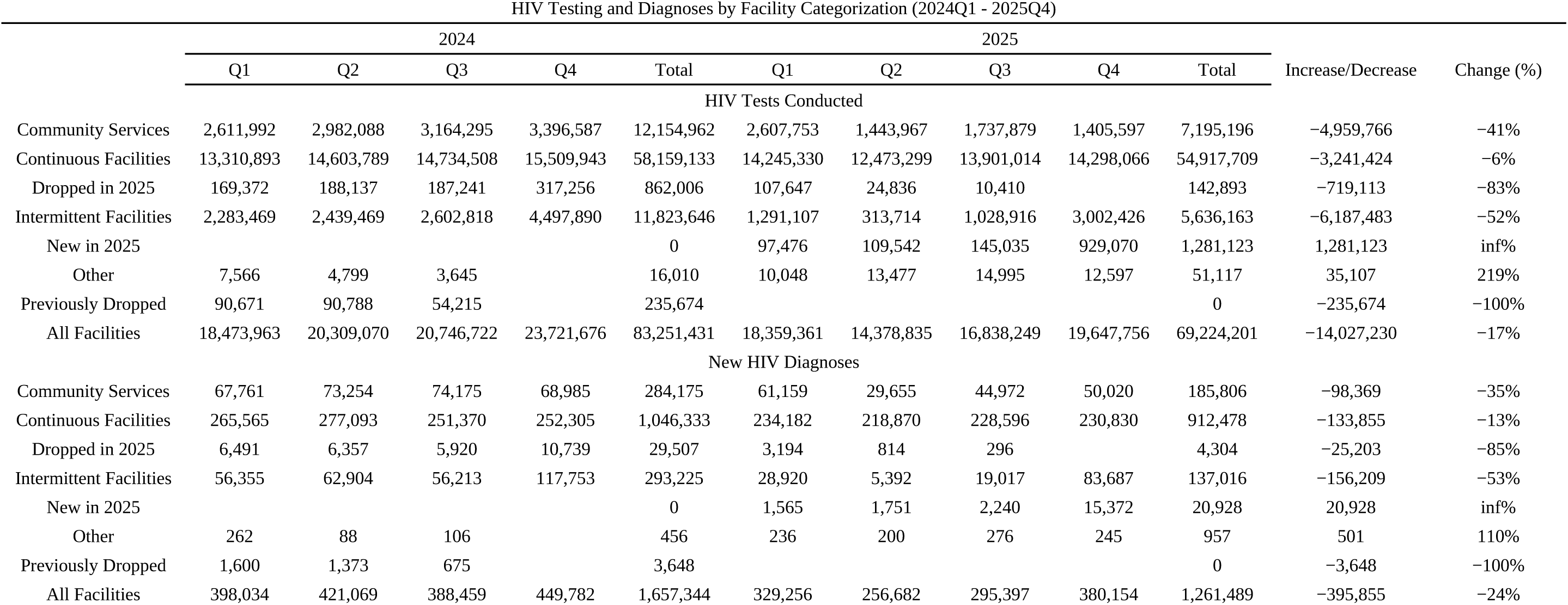
HIV Testing Indicators (2024Q1 - 2024Q4)

Declines in HIV diagnoses are most stark with a -13% reduction in continuous facilities (N=-133,855), -35% reduction in community services (n=-98,369), and a -29% in intermittent facilities on a Q4 to Q4 only basis (n=-34,066)

Concurrently, the number of people initiated on treatment declined substantially (Table 2). Initiations declined by -16% (n=-199,953) and -28% (n=-39,322) at continuous and intermittent facilities respectively.

Reduction in return to treatment (RTT) services were also observed with a -9% (n=-113,328) and -52% (n=-55,130) reduction from continuous and intermittent facilities.

### Impact on PMTCT service delivery

Overall, PMTCT services were also reduced, though more for infants than for pregnant women (Table 3). Testing and diagnoses among pregnant women remained relatively stable (-0.3% in testing, -1.7% in HIV diagnoses).

**Table 3:** PMTCT Indicators (2024Q1 - 2024Q4)

PMTCT and Early Infant Diagnoses and Treatment by Facility Categorization (2024Q1-2025Q4)
|  | 2024 |  |  |  |  | 2025 |  |  |  |  | Increase/<br>Decrease | Change (%) |
| --- | --- | --- | --- | --- | --- | --- | --- | --- | --- | --- | --- | --- |
|  | Q1 | Q2 | Q3 | Q4 | Total | Q1 | Q2 | Q3 | Q4 | Total |  |  |
| HIV Tests in PMTCT Programs |  |  |  |  |  |  |  |  |  |  |  |  |
| Continuous Facilities | 3,081,072 | 3,388,259 | 3,214,894 | 3,272,620 | 12,956,845 | 3,200,184 | 3,111,180 | 3,294,894 | 3,388,993 | 12,995,251 | 38,406 | 0% |
| Dropped in 2025 | 18,772 | 23,462 | 23,330 | 24,893 | 90,457 | 12,090 | 1,850 | 1,299 |  | 15,239 | -75,218 | -83% |
| Intermittent Facilities | 199,622 | 243,701 | 254,914 | 280,077 | 978,314 | 206,012 | 84,263 | 119,621 | 498,838 | 908,734 | -69,580 | -7% |
| New in 2025 |  |  |  |  | 0 | 26,361 | 32,181 | 34,187 | 46,032 | 138,761 | 138,761 | inf% |
| Other | 1,178 | 1,175 | 298 |  | 2,651 | 2,043 | 3,285 | 3,667 | 3,576 | 12,571 | 9,920 | 374% |
| Previously Dropped | 29,655 | 30,391 | 25,015 |  | 85,061 |  |  |  |  | 0 | -85,061 | -100% |
| All Facilities | 3,330,299 | 3,686,988 | 3,518,451 | 3,577,590 | 14,113,328 | 3,446,690 | 3,232,759 | 3,453,668 | 3,937,439 | 14,070,556 | -42,772 | -0% |
| New HIV Diagnoses in PMTCT Programs |  |  |  |  |  |  |  |  |  |  |  |  |
| Continuous Facilities | 123,862 | 133,343 | 125,297 | 127,302 | 509,804 | 122,549 | 122,719 | 119,792 | 117,859 | 482,919 | -26,885 | -5% |
| Dropped in 2025 | 646 | 857 | 707 | 677 | 2,887 | 340 | 67 | 77 |  | 484 | -2,403 | -83% |
| Intermittent Facilities | 23,797 | 27,244 | 24,536 | 25,538 | 101,115 | 11,596 | 1,701 | 9,258 | 93,944 | 116,499 | 15,384 | 15% |
| New in 2025 |  |  |  |  | 0 | 633 | 760 | 751 | 2,556 | 4,700 | 4,700 | inf% |
| Other | 119 | 48 | 15 |  | 182 | 46 | 52 | 78 | 74 | 250 | 68 | 37% |
| Previously Dropped | 637 | 508 | 242 |  | 1,387 |  |  |  |  | 0 | -1,387 | -100% |
| All Facilities | 149,061 | 162,000 | 150,797 | 153,517 | 615,375 | 135,164 | 125,299 | 129,956 | 214,433 | 604,852 | -10,523 | -2% |
| PBFW on Treatment |  |  |  |  |  |  |  |  |  |  |  |  |
| Continuous Facilities | 123,362 | 132,162 | 125,206 | 131,535 | 512,265 | 123,580 | 121,891 | 120,604 | 118,554 | 484,629 | -27,636 | -5% |
| Dropped in 2025 | 632 | 847 | 657 | 5,998 | 8,134 | 288 | 67 | 76 |  | 431 | -7,703 | -95% |
| Intermittent Facilities | 23,665 | 27,061 | 24,413 | 83,171 | 158,310 | 18,754 | 1,674 | 9,275 | 68,179 | 97,882 | -60,428 | -38% |

PMTCT and Early Infant Diagnoses and Treatment by Facility Categorization (2024Q1-2025Q4)
|  | 2024 |  |  |  |  | 2025 |  |  |  |  | Increase/<br>Decrease | Change (%) |
| --- | --- | --- | --- | --- | --- | --- | --- | --- | --- | --- | --- | --- |
|  | Q1 | Q2 | Q3 | Q4 | Total | Q1 | Q2 | Q3 | Q4 | Total |  |  |
| New in 2025 |  |  |  |  | 0 | 555 | 725 | 720 | 2,518 | 4,518 | 4,518 | inf% |
| Other | 47 | 42 | 14 |  | 103 | 49 | 49 | 77 | 71 | 246 | 143 | 139% |
| Previously Dropped | 637 | 514 | 226 |  | 1,377 |  |  |  |  | 0 | -1,377 | -100% |
| All Facilities | 148,343 | 160,626 | 150,516 | 220,704 | 680,189 | 143,226 | 124,406 | 130,752 | 189,322 | 587,706 | -92,483 | -14% |
| Early Infant HIV Diagnoses Testing |  |  |  |  |  |  |  |  |  |  |  |  |
| Community Services |  | 2 |  |  | 2 |  |  |  |  | 0 | -2 | -100% |
| Continuous Facilities | 125,176 | 127,981 | 127,585 | 127,256 | 507,998 | 119,135 | 114,420 | 124,151 | 120,582 | 478,288 | -29,710 | -6% |
| Dropped in 2025 | 1,388 | 1,339 | 1,309 | 1,021 | 5,057 | 302 | 50 | 60 |  | 412 | -4,645 | -92% |
| Intermittent Facilities | 37,390 | 37,438 | 37,196 | 33,276 | 145,300 | 22,407 | 1,124 | 11,257 | 13,315 | 48,103 | -97,197 | -67% |
| New in 2025 |  |  |  |  | 0 | 304 | 398 | 421 | 476 | 1,599 | 1,599 | inf% |
| Other | 18 | 512 | 549 |  | 1,079 | 542 | 524 | 553 | 580 | 2,199 | 1,120 | 104% |
| Previously Dropped | 324 | 339 | 165 |  | 828 |  |  |  |  | 0 | -828 | -100% |
| All Facilities | 164,296 | 167,611 | 166,804 | 161,553 | 660,264 | 142,690 | 116,516 | 136,442 | 134,953 | 530,601 | -129,663 | -20% |
| Infants Diagnosed HIV Positive |  |  |  |  |  |  |  |  |  |  |  |  |
| Continuous Facilities | 1,890 | 2,202 | 2,216 | 1,997 | 8,305 | 1,763 | 1,569 | 1,900 | 2,105 | 7,337 | -968 | -12% |
| Dropped in 2025 | 14 | 16 | 16 | 11 | 57 | 9 | 2 | 2 |  | 13 | -44 | -77% |
| Intermittent Facilities | 358 | 342 | 318 | 217 | 1,235 | 300 | 17 | 115 | 149 | 581 | -654 | -53% |
| New in 2025 |  |  |  |  | 0 | 3 | 5 | 12 | 17 | 37 | 37 | inf% |
| Other |  |  | 2 |  | 2 |  |  |  | 1 | 1 | -1 | -50% |
| Previously Dropped | 1 | 1 | 3 |  | 5 |  |  |  |  | 0 | -5 | -100% |
| All Facilities | 2,263 | 2,561 | 2,555 | 2,225 | 9,604 | 2,075 | 1,593 | 2,029 | 2,272 | 7,969 | -1,635 | -17% |
| Infants Diagnosed HIV Positive within 2 Months |  |  |  |  |  |  |  |  |  |  |  |  |
| Continuous Facilities | 854 | 1,000 | 1,057 | 941 | 3,852 | 779 | 650 | 756 | 950 | 3,135 | -717 | -19% |
| Dropped in 2025 | 9 | 6 | 8 | 3 | 26 | 3 |  | 2 |  | 5 | -21 | -81% |
| Intermittent Facilities | 180 | 181 | 171 | 104 | 636 | 129 | 8 | 55 | 73 | 265 | -371 | -58% |
| New in 2025 |  |  |  |  | 0 | 3 | 2 | 7 | 4 | 16 | 16 | inf% |
| Other |  |  | 2 |  | 2 |  |  |  | 1 | 1 | -1 | -50% |

PMTCT and Early Infant Diagnoses and Treatment by Facility Categorization (2024Q1-2025Q4)
|  | 2024 |  |  |  |  | 2025 |  |  |  |  | Increase/<br>Decrease | Change (%) |
| --- | --- | --- | --- | --- | --- | --- | --- | --- | --- | --- | --- | --- |
|  | Q1 | Q2 | Q3 | Q4 | Total | Q1 | Q2 | Q3 | Q4 | Total |  |  |
| Previously Dropped |  | 1 |  |  | 1 |  |  |  |  | 0 | -1 | -100% |
| All Facilities | 1,043 | 1,188 | 1,238 | 1,048 | 4,517 | 914 | 660 | 820 | 1,028 | 3,422 | -1,095 | -24% |
| Infants Placed on HIV Treatment |  |  |  |  |  |  |  |  |  |  |  |  |
| Continuous Facilities | 1,663 | 1,934 | 1,887 | 1,766 | 7,250 | 1,593 | 1,311 | 1,593 | 1,874 | 6,371 | -879 | -12% |
| Dropped in 2025 | 7 | 15 | 16 | 10 | 48 | 9 | 2 | 1 |  | 12 | -36 | -75% |
| Intermittent Facilities | 293 | 287 | 247 | 181 | 1,008 | 204 | 9 | 104 | 130 | 447 | -561 | -56% |
| New in 2025 |  |  |  |  | 0 | 2 | 5 | 9 | 14 | 30 | 30 | inf% |
| Other |  |  |  |  | 0 |  |  |  | 1 | 1 | 1 | inf% |
| Previously Dropped | 1 | 4 | 1 |  | 6 |  |  |  |  | 0 | -6 | -100% |
| All Facilities | 1,964 | 2,240 | 2,151 | 1,957 | 8,312 | 1,808 | 1,327 | 1,707 | 2,019 | 6,861 | -1,451 | -17% |

However, infant testing and diagnoses declined substantively. Infant testing at continuous facilities reduced testing by -29,710 (-6%) and diagnoses by -968 (-12%). Intermittent facilities, on a Q4 to Q4 basis saw a -60% reduction in testing (n=-19,961) and a -31% in diagnoses (n=-68). Infant treatment initiations declined inline with infant diagnoses reductions.

### Impact on PrEP

PrEP initiations declined across all facility types by -33%. At continuous facilities, there was a -27% decline in PrEP initiations (n=-569,312). and Q4 to Q4 PrEP initiations in intermittent facilities declined by -62%.

### Impact on human resources for health

The PEPFAR-supported workforce declined substantially between 2024 and 2025 (Figure 4). Total staff employed was reduced by -22% (n=-76,051) with a -28% reduction in full-time equivalent staff (n=-69,528). By healthcare worker (HCW) category, the number of ancillary HCWs declined by -34% (n=-69,602) while there were 12% more clinical workers (n=6,992).

**Figure 4:**
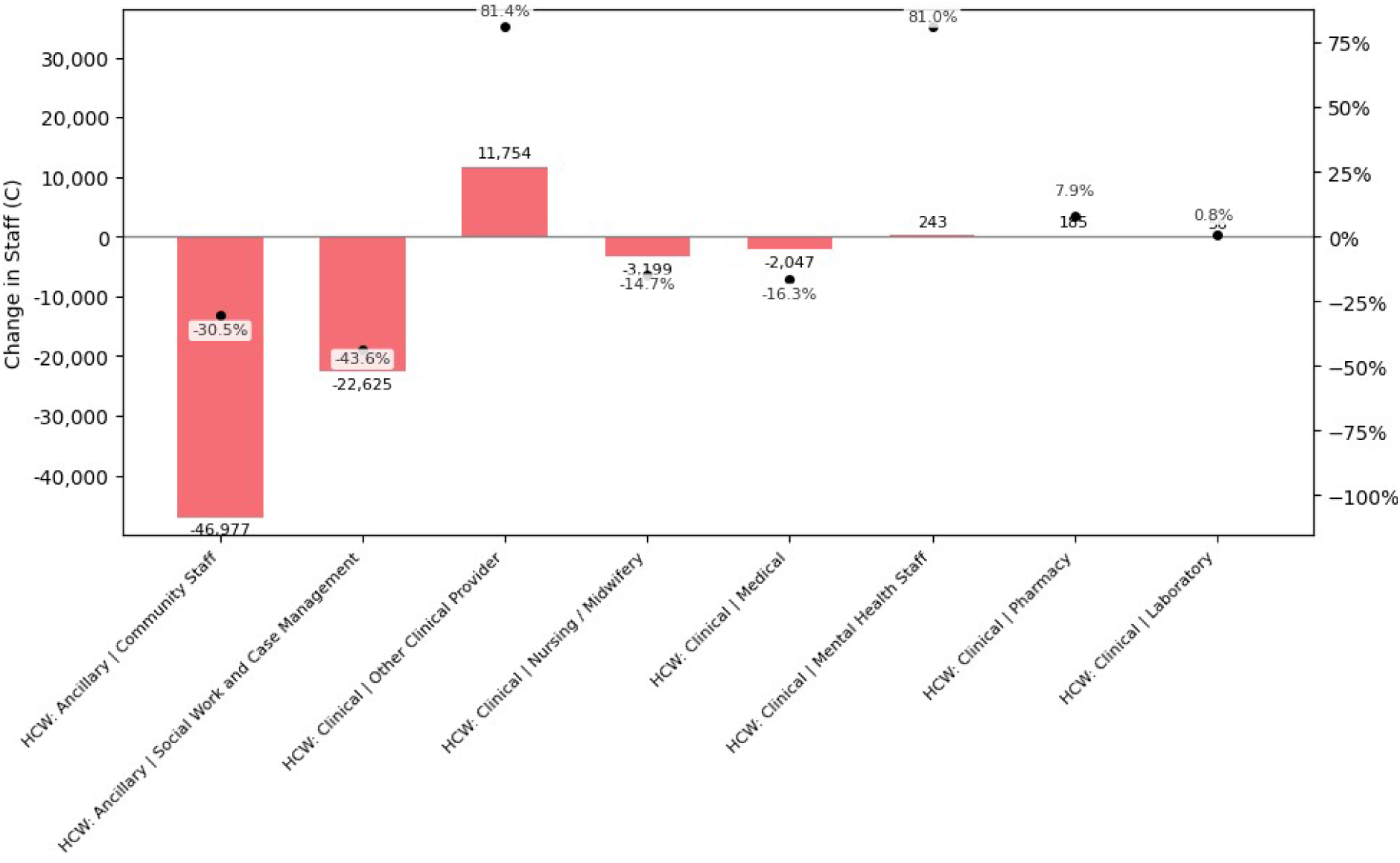
Change in employed staff by HCW category and cadre (2024 - 2025)

Other cadres decreased by -16% (n=-13,441). Overall combined ancillary and clinical HCWs employed declined by -24% (n=-62,541).

Country-level patterns can be seen in Supplement 2.

## Discussion

The levels of disruption seen in the PEPFAR 2025 results reveal both the resilience of healthcare systems in maintaining treatment continuity and raise serious concerns for the prospects of achieving and maintaining epidemic control. Interpreting these results requires careful attention to the reporting disruptions that accompanied the foreign aid freeze, which this analysis addresses using the unofficial Q1-Q3 data to categorize facilities according to their exposure to disruptions and to evaluate the lasting impact of the foreign aid freeze on actual service provision.

That overall numbers of people on treatment remain relatively stable at the PEPFAR-wide level or even country level obscures substantial changes occurring beneath the aggregated results. That Continuous Facilities successfully rebounded to slightly above their 2024 level is consistent with these facilities largely maintaining at least some level of support from PEPFAR and are primarily owned and operated by ministries of health. Approximately 75% of all facilities in which PEPFAR reported data in 2024 were owned by ministries of health or other government agencies.[11] Access to treatment is also a lagging indicator of the overall health system performance as patients stable on treatment already have strong routines in place for continually collecting medication. The lack of geographic overlap between where sites were dropped and new sites added, combined with net declines in treatment numbers in affected geographies, suggests that patients from dropped sites were largely not transitioned to other facilities, but rather that already established clinics were absorbed into PEPFAR reporting and consist of individuals who were already on treatment outside of PEPFAR support, not as a result of it.

The significant declines in HIV testing, diagnoses, treatment initiations, and treatment retention programming, however, raise serious concerns for countries’ capacity to maintain progress toward the 95-95-95 targets. Drops in HIV diagnoses of -13% in Continuous Facilities, -35% from Community Services, and -29% in Intermittent Facilities (based on Q4-Q4 comparisons), jeopardize progress toward the first or second 95. While these are correctly described as “HIV Diagnoses” rather than “New HIV Diagnoses” as they are inclusive of individuals who have been previously diagnosed re-testing for HIV, re-testing remains - in most countries - a vital pathway for patients to re-engage and re-initiate treatment after a treatment interruption. Treatment initiations dropped by rates similar to HIV diagnoses and growth of people accessing treatment was down 48.6% in Continuous Facilities and -1.7% in Intermittent Facilities (Q1-Q4).

The relatively minor-seeming reductions of -1.7% in people on treatment in Intermittent Facilities relative to 0.5% growth in Continuous Facilities should not be dismissed. Intermittent Facilities are more likely to represent where PEPFAR implementing partners support has been reduced or withdrawn, taking with it the retention and wrap-around programming that keeps patients engaged in care. Sustained loses of this kind, even at small scales, risks progressive declines in community viral suppression that open the possibility for HIV resurgence to occur.

The decline in PMTCT infant testing, diagnoses, and treatment initiations is particularly concerning given the poor survival rate of infants who contract HIV at birth or during breastfeeding when they do not access treatment. The -6% in testing and -12% in diagnoses in Continuous Facilities and -60% and -31% in Intermittent Facilities (Q4-Q4), respectively strongly indicate that PMTCT programs have been substantially impacted by the disruptions of 2025.

Details on prevention programming in the facility level data are limited, though the scope of restrictions suggest substantial reductions. . The limited waiver for PEPFAR to the foreign assistance freeze in February 2025 did not include most prevention programming, including voluntary medical male circumcision (VMMC), gender-based violence (GBV) programming, key and priority population programming, and restricted PrEP only to pregnant and breastfeeding women.[19] Additionally, condom and lube procurement has seen significant disruptions.[20] In 2024, PEPFAR supported VMMC for 2.5 million boys and men, and intervention which reduces their risk of acquiring HIV by 60%.[21,22] While interruptions in VMMC are unlikely to affect incidence immediately, they represent a meaningful long term risk of HIV resurgence. The restrictions on PrEP programming - despite the U.S. government’s interest in lenacapavir - undermine near-term prevention efforts..

On HRH, the small increase in clinical HCWs was almost entirely a product of increases in the “Other Clinical Provider” cadre (n=11,754, +81%), which is primarily made up of case managers, social workers, and lay testing counselors. These increases in many cases are likely to include significant re-classifications from the “Social Work and Case Management” cadre of ancillary HCW (n=-22,625, -44%) rather than a strategic intention to re-allocate resources. This can be seen in the data from Cameroon and Malawi. Some countries such as Mozambique and Nigeria did show large increases in HRH.

The AFGHS explicitly committed to maintaining 100% of frontline health worker costs in FY2026 currently supported by the U.S. government, citing the 270,000 individuals supported by PEPFAR funding, including 208,800 Community Health Workers and 22,700 nurses.[23] These numbers are consistent with the 2024 HRH data that had 205,726 Ancillary HCW Staff and 21,819 Nurses and Midwives. Both of these have been reduced substantially by -69,602 (-34%) and -3,199 (-15%), respectively. Total direct service delivery HCWs have been reduced from 264,422 to 201,881 (-24%) in direct contradiction of the AFGHS’ written commitment.

### Strengths and Weaknessess

This analysis methodology accounts for the incompleteness in PEPFAR reported data to hold a consistent comparison of results across the time period and distinguishing reporting disruptions from actual service delivery changes. PEPFAR data are a global data set but are not necessarily representative at the national level of all service delivery in the country. Facility level classifications are not necessarily perfect and there could be some residual under-reporting of data for specific indicators.

## Conclusion

This is potentially the last data set PEPFAR will ever release. The AFGHS and the Bureau of Global Health Security and Diplomacy (GHSD) have stated an intention to replace PEPFAR’s comprehensive data system (DATIM) with reporting results out of national data systems untethered to actual PEPFAR support or funding.[24] Under the terms of the memoranda of understanding (MOUs) that the U.S. State Department is currently signing with partner governments, public disclosure of data from those data sets is prohibited, including with external researchers, academics, or advocacy organizations. Moreover, those data sets are likely to omit any significantly disaggregated age and sex data and will only focus on an extremely limited set of indicators established in the MOUs - namely: the total number of people on treatment; total number of HIV diagnoses (infants and anyone over 18 months of age separately); and the percent of HIV-positive pregnant and breastfeeding women who are on ART.[24] Without public data sets that enable national, sub-national, facility-level, and implementing mechanism level scrutiny, it is virtually impossible for external oversight and accountability to take place, whether within the U.S. government or outside it.

These findings represent a troubling inflection point for what has been widely cited as the most transparent and effective global health program in history and credited with saving more than 26 million lives. The dismantling of the data systems and accountability structures that made PEPFAR uniquely effective risks compounding the direct service losses documented here. Most concerningly, the collapse of this programming is likely to be felt in the coming years as 95-95-95 slips to 90-90-95 and below and HIV incidence (never truly curtailed to the extent necessary) increases again. The U.S. Congress retains the authority and capacity to intervene to ensure global health programming is adequately funded and effective, but to do so it must take significant steps to override the dismantling of this public and accountable program.

## Data Availability

Data are from publicly available sources

https://data.pepfar.gov

